# Midwifery and nurse staffing of inpatient maternity services – a systematic scoping review of associations with outcomes and quality of care

**DOI:** 10.1101/2021.03.27.21254457

**Authors:** Lesley Turner, Peter Griffiths, Ellen Kitson-Reynolds

**Author notes:** Co-authors (Peter Griffiths, Ellen Kitson-Reynolds). **CRediT roles** Lesley Turner Conceptualisation, Methodology, Investigation, Writing – original draft Professor Peter Griffiths Conceptualisation, Methodology, Writing – review and editing Dr Ellen Kitson-Reynolds Validation, Writing – review and editing.

## Abstract

**Objective:** To undertake a scoping literature review of studies examining the quantitative association between staffing levels and outcomes for mothers, neonates, and staff. The purpose was to understand the strength of the available evidence, the direction of effects, and to highlight gaps for future research.

**Data Sources:** Systematic searches were conducted in Medline (Ovid), Embase (Ovid), CINAHL (EBCSCO), Cochrane Library, TRIP, Web of Science and Scopus.

**Study Selection and Review methods:** To be eligible, staffing levels had to be quantified for in-patient settings, such as ante-natal, labour/delivery or post-natal care. Staff groups include registered midwives, nurse midwives or equivalent, and assistant staff working under the supervision of registered professionals. Studies of the quality of care, patient outcomes and staff outcomes were included. All quantitative designs were included, including controlled trials, time series, cross-sectional, cohort studies and case controlled studies.

Data were extracted and sources of bias identified by considering the study design, measurement of exposure and outcomes, and risk adjustment. Studies were grouped by outcome noting the direction and significance of effects.

**Results:** The search yielded a total of 3280 records and 21 studies were included in this review. There were three randomised controlled trials, eleven cohort studies, one case control study and six cross sectional studies. Seventeen were multicentre studies and nine of them had over 30,000 participants.

Reduced incidence of epidural use, augmentation, perineal damage at birth, postpartum haemorrhage, maternal readmission, and neonatal resuscitation were associated with increased midwifery staff. Few studies have suggested a negative impact of increasing staffing rates, although a number of studies have found no significant differences in outcomes. Impact on the mode of birth were unclear. Increasing midwifery support staff was not associated with improved patient outcomes. No studies were found on the impact of low staffing levels for the midwifery workforce.

**Conclusions and Implications for practice:** Although there is some evidence that higher midwifery staffing is associated with improved outcomes, current research is insufficient to inform service planning. Studies mainly reported outcomes relating to labour, highlighting a gap in research evidence for the antenatal and postnatal periods. Further studies are needed to assess the costs and consequences of variations in maternity staffing, including the deployment of maternity care assistants and other staff groups.

## Introduction

Inpatient maternity services provide antenatal, intrapartum, and postnatal care for women and babies with additional needs, and for those choosing to give birth in a hospital environment. There is much variation in the staffing levels for these in-patient units (Zbiri et al., 2018; Zhou et al., 2019; NHS Digital, 2020). Maternity professionals have concerns about low staffing levels and report that this poses a threat to safety (Smith et al., 2009; Simpson et al., 2016). Staffing levels have been implicated in a number of near-miss cases and sub-optimal outcomes (Ashcroft et al., 2003). Problems with inadequate staffing were identified in over a quarter of stillbirths reported in the UK from 2015-2017 (Manktelow et al., 2017).

In order to inform workforce planning, managers need evidence based guidelines to inform their staffing decisions. One such guideline from the UK is the recommendation that women should receive dedicated care from one midwife during labour (Royal College of Obstetricians and Gynaecologists, 2007). Guidelines differ in other parts of the world, and California was one of the first states to mandate a staffing ratio of no more than 2 patients in active labour to 1 nurse (Coffman et al., 2002). Evidence underpinning this was sparse at this time, although a later Cochrane review confirmed that continuous support in labour (from hospital staff or birth supporters) was associated with a higher rate of vaginal birth, reduced caesarean section, reduced instrumental birth and improved Apgar scores (Hodnett et al., 2013; Bohren et al., 2017). A large number of women still receive labour care by a core team of midwives, who also deliver care in the antenatal and postnatal wards of the hospital (Care Quality Commission, 2020). This is despite mounting evidence to support the roll out of continuity of carer (Sandall et al., 2016) which offers benefits in terms of reduced rates of stillbirth, premature births and medical interventions.

The relationship between staffing and outcomes is important in determining the level at which harm can occur or the level at which there is no additional tangible benefit in deploying more midwives. This is important as cost-effectiveness must be considered due to the scarcity of resources and competing demands in health care. Poor staffing has been implicated in a number of error reviews and reports of near-misses in maternity care (Ashcroft et al., 2003; Karimi et al., 2016). The cost of litigation in maternity care is soaring (Tingle, 2016), and the human cost of poor outcomes is immeasurable. Complexity in maternity cases is increasing, so there is likely to be sustained demand for complex inpatient maternity care, requiring the expertise of core staff in these areas.

The impact of inadequate staffing is far-reaching and midwives have reported on the areas that have been missed due to high workload or time constraints (Simpson et al., 2016; Simpson et al., 2017; Haftu et al., 2019). This includes measuring vital signs, medicines administration, noting changes in acuity, response in emergencies and emotional support (Bick et al., 2014; Simpson et al., 2016). This can lead to reduced opportunities to identify deterioration and to rescue from preventable patient harm, such as fetal demise in labour, neonatal hypoglycaemia or infection (Simpson et al., 2017). One outcome that may be sensitive to staffing is the rate of term babies admitted to the neonatal unit (Clapp et al., 2019), causing separation from mothers and great cost to the health service.

A large body of evidence exists within nursing to suggest that a number of outcomes are sensitive to changes in staffing, such as falls, pressure ulcers and mortality (Patrician et al., 2011; Staggs et al., 2012; Griffiths et al., 2018). In an observational study of over 422,000 surgical patients in Europe, the increase in nurses workload by one patient increased the risk of a patient dying within 30 days by 7% (Aiken et al., 2014). There have been fewer studies in the midwifery literature, although a significant review was conducted by Bazian (2015) which summarised evidence from eight studies and highlighted a number of gaps in the research evidence. They found that most studies related to labour outcomes and mode of birth, although there was no consensus on the direction of effects for most maternal and fetal outcomes. This area is worthy of further exploration as a number of new studies have been published since the Bazian review (Bazian, 2015). Before future research is commissioned it is important to review the studies to date, and to establish what is known (and unknown) about the relationship between staffing and patient outcomes.

A further driver for interest in this area is the training and development of maternity support workers. Their role provides the opportunity for task-shifting and complementing the work of midwives. It is unclear whether the evidence supports the widespread development of these roles, although an evaluation by Griffin et al. (2012) suggests a potential positive impact on breastfeeding, parent education and discharge procedures. Preliminary work has been undertaken on the economics of skill mix in maternity care by Cookson et al. (2014) and Laliotis et al. (2018), although there is concern about the quality of data on effectiveness, due to the use of aggregate data to measure staffing and the potential for unmeasured confounding in observational studies.

## Methods

The aim of this scoping literature review was to identify and summarise studies which examine the association between staffing levels of registered midwives and the outcomes for mothers and neonates. The purpose was to examine the strength of the available evidence, the direction of effects, and to highlight gaps for future research.

The review addressed the following specific questions.

What is the extent and nature of the body of knowledge relating midwifery staffing to outcomes, in terms of the number of studies, designs, methodology, participants, settings and outcomes investigated?

Is there an association between the midwifery staffing levels for in-patient services and outcomes and quality of care, and do outcomes differ when the proportion of registered staff to support workers varies?

### Design

A scoping literature review methodology was selected in order to summarize the breadth of the evidence from a range of sources (Levac et al., 2010). Unlike a systematic review, a scoping review allows researchers to identify all the relevant literature regardless of study design. A protocol was not registered in advance as this scoping review developed iteratively to discover the nature of the literature available.

### Search strategy

Searches were completed in Medline (Ovid), Embase (Ovid), CINAHL (EBCSCO), Cochrane Library, TRIP, Web of Science and Scopus on 6 th April 2020. Search terms were entered as key words and subject headings, to identify primary research relating to staffing and maternity care (See Appendix 1 for full search strategy). No limitations were placed on the date of publication.

The reference lists of eligible studies were scanned to identify further references. All eligible studies were entered into the Cited Reference Search in Web of Science to identify citations and potential new primary studies in the same field.

### Study selection

Studies were eligible for inclusion if they investigated the quantitative association between a measure of midwifery staffing levels and/or skill mix and outcome for mother baby, or staff members, costs or quality of care. All quantitative designs were included including controlled trials, time series, cross-sectional, cohort studies and case controlled studies. Studies on the effects of implementing changes to staffing levels or mix were included, as were studies on the effects of implementing a mandatory minimum staffing policy or a tool to measure demand and guide staffing decisions.

To be eligible for inclusion, staffing levels had to be quantified in measures such as staff per bed, staff to mother ratio, or hours per patient day. An assumption was made that continuous support from a midwife in labour was similar to a staffing ratio of 1:1, and therefore papers reporting staffing in this way were eligible for inclusion. Staff groups include registered midwives, nurse midwives or equivalent, and assistant staff working under the supervision of registered professionals. Studies reporting a quantitative measure of subjective staffing adequacy were included but purely qualitative studies were excluded.

Staffing in any or all inpatient settings were considered including ante-natal, labour/delivery and post-natal care. Studies which were based in neonatal units and midwifery community settings were excluded.

All references arising from the search were imported into Endnote X9™ reference management software where duplicates were removed. Studies were screened and excluded if titles were unrelated to the subject area. The abstracts of 266 studies were read and studies excluded if it was clear that the inclusion criteria were not met by reading the abstract alone. Forty-six full text articles were screened against the inclusion criteria. All included papers were checked, and the decision verified by at least two reviewers. Of the excluded papers, double rating of a sample suggested a high level of agreement. Data charting was performed by one investigator.

Statistical meta-analysis was not attempted but all results were tabulated to show both the direction and statistical significance of the observed effects. From this a description of the overall pattern of results was derived. Sources of bias were identified by considering the study design, measurement of exposure and outcomes, and risk adjustment.

## Results

### Summary of included studies

The online searches yielded a total of 3280 records. The PRISMA flow diagram is shown in Figure 1 below. Twenty one separate studies were identified which were published from 1988 −2020. These studies are tabulated in detail in Appendix 2. Data were extracted from 23 papers as two studies were reported separately. One study was available as an abstract only (Mercer, 2016). There were three randomised controlled trials, eleven cohort studies, one case control study and six cross-sectional studies.

**Figure 1:**
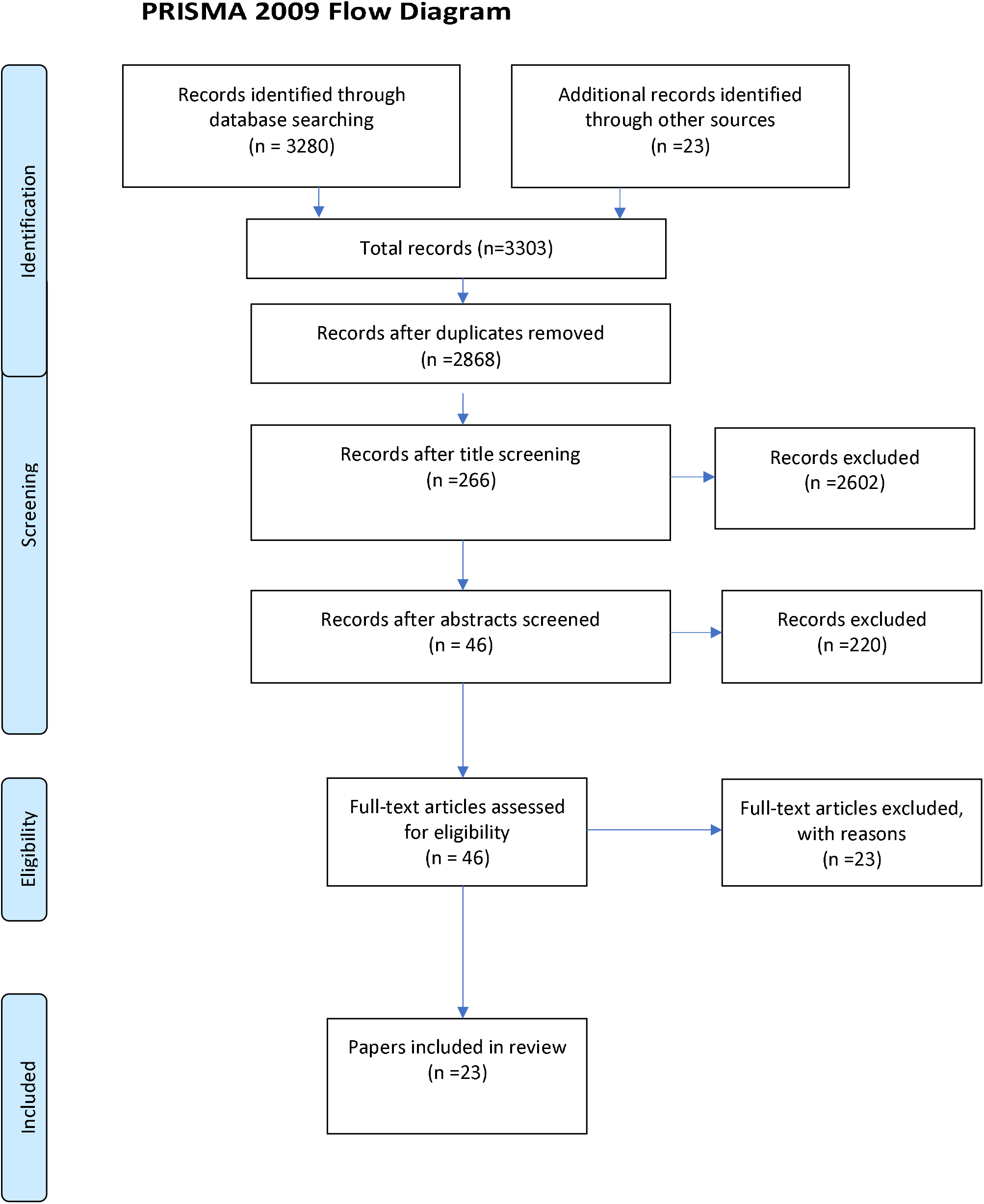
Outcome of search strategy

Nine studies were conducted in the UK, and the remaining studies were conducted in USA, Canada, France, Italy, Indonesia, Korea, Thailand and Iran. Six studies included only participants at low risk of complications. Three studies included only complex cases such as women having postpartum haemorrhage (Prapawichar et al., 2020), those having oxytocin in labour (Clark et al., 2014) or caesarean section (Kim et al., 2016). The majority of studies (14/21) reported only outcomes relating to labour and birth. No studies of antenatal inpatient care were found, and there were four studies of postnatal care outcomes, including those studying readmission rates.

There were 17 multicentre studies and many were large. Nine studies had over 30,000 participants and five studies had over 400,000 participants. In terms of measurement of staffing, 16 studies used the term ‘midwife’ while others looked at staffing by ‘nurses’ or ‘nurse-midwives’ in a labour setting. Three studies also included the impact of health care assistant/support worker staffing, and eight studies also examined medical staffing in terms of obstetricians, anaesthetists or neonatal doctors.

### Quality of the evidence

Three randomised controlled trials (Gagnon et al., 1997; Hodnett et al., 2002; Kashanian et al., 2010) compared patients all receiving one to one care in labour with usual staffing levels, although all had some limitations. Hodnett et al. (2002) excluded patients where one-to-one care was deemed medically necessary. Kashanian et al. (2010) included only 100 women and the usual labour care involved a lack of privacy, no birth companion and women were not permitted to eat and drink. The third RCT (Gagnon et al., 1997) was relatively small and incorporated other therapeutic measures along with the one-to-one care which limits the ability to assess the effects of the staffing ratio alone.

Of the eleven cohort studies, only the Tucker et al. (2003) study provided data on objective patient outcomes while also adjusting for baseline risk and other confounders. Other cohort studies considered care processes such as time to theatre transfer for caesarean section, quality of record keeping, mode of birth or labour interventions (Cerbinskaite et al., 2011; Knape et al., 2014; Rowe et al., 2014; Bailey et al., 2015; Zbiri et al., 2018). These outcomes may not translate directly into benefits for patients. The study by Clark et al. (2014) was conducted in a select patient group receiving oxytocin, limiting the generalisability of findings. The measurement of staffing was based on opinion, and the background risk was not adjusted for. The Dani et al. (2020) study did not measure staffing exposure directly and was at risk of bias due to differences in settings and patient acuity between the two groups. Cohort studies by Kim et al. (2016) and Stilwell et al. (1988) were deemed to be at high risk of bias in the assessment of staffing exposure and had limited risk adjustment. Mercer (2016) was published only as an abstract and therefore the methodology could not be scrutinised.

Of the six cross-sectional studies, four were large scale studies which used routine data to assess exposure to staffing and patient-centred outcomes such as perineal damage, maternal mortality, readmission rates, still birth and neonatal mortality (Joyce et al., 2004; Gerova et al., 2010; Sandall et al., 2014; Makhfudli et al., 2020). Other cross sectional studies focused on the outcome of mode of birth (Joyce et al., 2002; Gerova, 2014) or had a narrow focus on epidural use (Kpéa et al., 2015). All of these studies controlled for risk in terms of maternal age, deprivation, and some measures of clinical risk. These cross-sectional studies considered aggregate measures of staffing such as the number of midwives employed at institutional level or the number of midwives in relation to patients or births. This represents a major difficulty in determining that staffing exposure is causally linked to outcomes for patients, as the time period and fluctuating staffing exposure may not match patient stay. It also does not account for deployment of midwives within the service as some may have non-clinical roles.

### Maternal outcomes in relation to staffing

Nine studies examined the outcomes for mothers after birth (Table 1). On the whole, most of these suggest improved outcomes where more staff were present. The outcomes studied included severe maternal outcome (death or near miss), perineal trauma, post-partum haemorrhage, maternal readmission, satisfaction, and maternal infection.

**Table 1:**
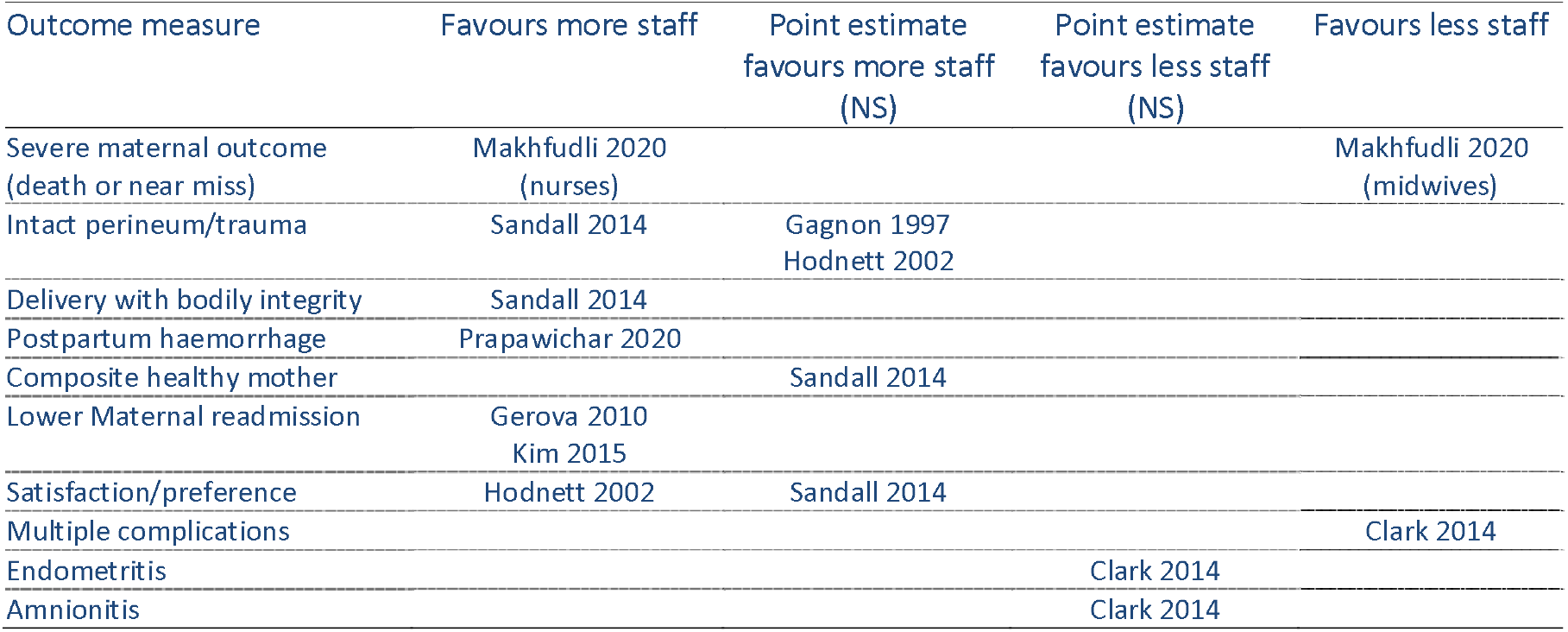
Maternal outcomes in relation to staffing

| Outcome measure | Favours more staff | Point estimate<br>favours more staff<br>(NS) | Point estimate<br>favours less staff<br>(NS) | Favours less staff |
| --- | --- | --- | --- | --- |
| Severe maternal outcome<br>(death or near miss) | Makhfudli 2020<br>(nurses) |  |  | Makhfudli 2020<br>(midwives) |
| Intact perineum/trauma | Sandall 2014 | Gagnon 1997<br>Hodnett 2002 |  |  |
| Delivery with bodily integrity | Sandall 2014 |  |  |  |
| Postpartum haemorrhage | Prapawichar 2020 |  |  |  |
| Composite healthy mother |  | Sandall 2014 |  |  |
| Lower Maternal readmission | Gerova 2010<br>Kim 2015 |  |  |  |
| Satisfaction/preference | Hodnett 2002 | Sandall 2014 |  |  |
| Multiple complications |  |  |  | Clark 2014 |
| Endometritis |  |  | Clark 2014 |  |
| Amnionitis |  |  | Clark 2014 |  |

Delivery with bodily integrity and intact perineum were more common when more midwives were employed (Sandall et al., 2014). This finding of reduced perineal trauma was supported by studies by Gagnon et al. (1997) and Hodnett et al. (2002) although significance was not reached. In the case control study by Prapawichar et al. (2020), hospitals which had below the standard nurse midwife to patient ratio had significantly increased odds of postpartum haemorrhage OR 2.3 (95% CI 1.08 to 4.92, p=0.03). Two studies found that maternal readmission was lower when more midwives or nurses were employed in the organisation (Gerova 2010, Kim 2015).

In contrast to this, the study by Clark et al. (2014) found opposite effects for rates of complications in their population of high risk women receiving oxytocin. The lack of risk adjustment in this study could not eliminate confounding by indication, that is higher risk women had higher staffing levels because of the increased risk. Makhfudli et al. (2020) found that the odds of a severe maternal outcome, as defined by World Health Organization (2019) was lower when women were admitted to units with higher nursing staffing (OR 0.48, 95% CI 0.31 to 0.74) but rates were increased in units where midwifery staffing was higher (OR 1.81, 95% (CI 1.07 to 3.06).

### Neonatal outcomes in relation to staffing

Ten studies examined the outcomes for neonates (Table 2). Outcomes studied included Apgar scores, birth asphyxia, need for neonatal resuscitation, breastfeeding, admission to the neonatal unit, stillbirth, neonatal death and a composite measure entitled healthy baby. Other potentially important outcomes for babies including neonatal readmission, neonatal hypoglycaemia, sustained breastfeeding, jaundice, and weight loss were not studied.

**Table 2:** Neonatal outcomes in relation to staffing

| Outcome measure | Favours more staff | Point estimate favours more staff (NS) | No difference or no data on direction | Point estimate favours less staff (NS) | Favours less staff |
| --- | --- | --- | --- | --- | --- |
| Apgar score |  | Tucker 2003<br>Kashanian 2010 |  |  | Gagnon 1997 |
| Lower Birth asphyxia |  | Clark 2014 |  | Hodnett 2002 |  |
| Lower rates Neonatal resus | Hodnett 2002 | Tucker 2003 |  |  |  |
| Lower rates Neonatal resus (excluding bag/mask only) | Tucker 2003 |  |  |  |  |
| Lower Stillbirth |  |  | Joyce 2004 |  |  |
| Lower Neonatal death |  |  | Joyce 2004<br>Stilwell1998 |  |  |
| Composite healthy baby |  | Sandall 2014 |  |  |  |
| Exclusive breastfeeding | Dani 2020 |  |  |  |  |
| Admission to Neonatal unit | Dani 2020 | Hodnett 2002<br>Tucker 2003 |  | Gagnon 1997 |  |
| Neonatal length of stay |  |  |  | Hodnett 2002 |  |
| Perinatal complications |  |  | Mercer 2016 |  |  |

Three studies report significantly improved outcomes which favour more staff, and one study shows results in the opposite direction. Dani et al. (2020) found higher breastfeeding rates with increased staffing (88% vs 78%, p=0.048), although comparisons took place in two different settings. They also report lower Neonatal Unit admission (2% vs 9%), and this is supported by further studies by Hodnett et al. (2002) and Tucker et al. (2003), although these findings did not reach significance. Gagnon (1997) provides evidence to the contrary, with rates of neonatal unit admission of 7.2% vs 4.9%, RR1.46 (95% CI 0.67, 3.18), thereby presenting a mixed picture for this outcome. Considering the overall pattern, 11 studies have point estimates in favour of more staff while four show results favouring less staff.

Of the higher quality studies (Tucker et al., 2003; Sandall et al., 2014), these suggest that higher staffing was associated with improved neonatal outcomes. Tucker et al. (2003) reported that fewer babies needed neonatal resuscitation using advanced measures (OR 0.97, 95% CI 0.94, 0.99). This was also noted by Hodnett et al. (2002) although no risk adjustment was undertaken in this study.

### Events during labour

Ten studies examined events during labour in relation to staffing (Table 3). Outcomes studied included the quality of record keeping, continuous fetal monitoring in low risk women, fetal distress, augmentation of labour, epidural use, speed of theatre transfer for caesarean section, and length of labour. These care process measures are difficult to interpret as they may not translate into differences in patient outcomes. Many of the findings favour more staff, with seven comparisons reaching statistical significance in that direction. Ten further comparisons show non-significant results in favour of more staff. Three comparisons favour having less staff, although some of these result from subgroup analyses.

**Table 3.** Events during labour in relation to staffing

| Outcome measure | Favours more staff | Point estimate favours more staff (NS) | Point estimate favours less staff (NS) | Favours less staff |
| --- | --- | --- | --- | --- |
| Completeness of partogram | Bailey 2015 (hrs 0-8 of shift) | Bailey 2015 (hrs 8-12 of shift) |  |  |
| Completeness of note keeping |  | Bailey 2015 (hrs 0-8 of shift) | Bailey 2015 (hrs 8-12 of shift) |  |
| Continuous fetal monitoring | Hodnett 2002 |  |  |  |
| Appropriate fetal monitoring |  | Tucker 2003 low risk women | Tucker 2003 high risk women |  |
| Less Fetal distress | Clark 2014 |  |  |  |
| Less oxytocin use / augmentation | Rowe 2014 in multiparous | Gagnon 1997<br>Kashanian 2010<br>Rowe 2014 in primiparous |  |  |
| Time to delivery interval for c-section | Cerbinskaite 2011 |  |  |  |
| Less Epidural use | Kpéa 2015 | Gagnon 1997<br>Joyce 2002<br>Rowe 2014 in nulliparous |  |  |
| Shorter Length of labour | Kashanian 2010 | Gagnon 1997 |  |  |

Fetal distress was lower in facilities that offered 1:1 care more frequently (Clark et al., 2014) and the completeness of the partogram improved (Bailey et al., 2015). Kpéa et al. (2015) found that if the midwifery workload was high, 58.3% of women had an epidural or spinal for pain relief, compared to 49.7% if the workload was not high (OR 1.1, 95% CI 1.0-1.2). This finding was also supported by other studies, although non-significant effects were seen (Gagnon et al., 1997; Joyce et al., 2002; Rowe et al., 2014). Lower staffing was associated with higher augmentation rates, and this reached significance for multiparous women (Rowe et al., 2014). These findings suggest higher intervention rates when staffing levels fall, possibly representing a lack of support for women to manage pain or to facilitate progress of labour.

Cerbinskaite et al. (2011)studied the time taken to enter theatre for emergency caesarean section, and found this to be reduced when more midwives were present. For example, transfer time to theatre for grade 1 caesarean section was achieved within 15 mins for 81/82 (99%) cases where staffing was 1:1 or better, compared to 34/40 (85%) when the ratio fell below this target.

### Mode of birth in relation to staffing

Ten studies examined mode of birth as an outcome measure, examining rates of emergency caesarean section, instrumental birth and spontaneous vaginal birth (Table 4). The results were mixed, and no patterns emerged favouring more or less staff.

**Table 4.** Mode of birth in relation to staffing

| Outcome measure | Favours more staff | Point estimate favours more staff (NS) | Point estimate favours less staff (NS) | Favours less staff |
| --- | --- | --- | --- | --- |
| Lower Caesarean birth rate | Kashanian 2010<br>Zbiri 2018<br>(elective cs) | Clark 2014<br>Gagnon 1997<br>Hodnett 2002<br>Joyce 2002<br>Sandall 2014<br>(emergency) | Gerova 2014<br>Rowe 2014 in<br>multiparous<br>Sandall 2014 (elective)<br>Zbiri 2018<br>(urgent or intrapartum cs) | Rowe 2014 in<br>nulliparous |
| Lower Instrumental birth |  | Joyce 2002<br>Hodnett 2002<br>Rowe 2014 in<br>nulliparous<br>Knape 2014* | Gagnon 1997<br>Gerova 2014<br>Rowe 2014 in<br>multiparous |  |
| Increased Spontaneous vaginal birth / Normal birth | Gerova 2014 | Sandall 2014<br>Rowe 2014 in<br>nulliparous | Hodnett 2002<br>Rowe 2014 in<br>multiparous |  |
| Increased Straightforward birth |  |  | Rowe 2014 in<br>nulliparous | Rowe 2014 in<br>multiparous |

\*Knape (2014) studied lower caesarean section or operative birth as one outcome

Measures of birth without assistance were defined differently in the studies, using the terminology ‘normal birth’ and ‘spontaneous vaginal birth’ at times. Within this theme, only Gerova (2014) found a significant association between increased staffing and more normal birth, while studies by Sandall (2014), Hodnett (2002) and Rowe (2014) offered inconclusive findings. An extension of this outcome ‘straightforward birth’ was used by Rowe (2014) to include unassisted birth with no serious perineal trauma or blood transfusion.

In terms of caesarean section rates, only two studies (Kashanian et al., 2010; Zbiri et al., 2018) found a positive association between more staff and reduced caesarean section rate. Rowe et al. (2014) found the opposite, in that understaffing was significantly associated with reduced caesarean section rates, and this was significant for nulliparous women. The majority of other studies examining this outcome found no significant differences (Gagnon et al., 1997; Hodnett et al., 2002; Joyce et al., 2002; Clark et al., 2014; Gerova, 2014; Sandall et al., 2014; Kim et al., 2016). All studies examining the effect of staffing on instrumental birth had non-significant findings and the directions of effect were not consistent (Joyce 2002, Gagnon 1997, Gerova 2014, Hodnett 2002, Rowe 2014).

### Effect of midwifery support worker staffing

Three studies (Gerova 2014, Sandall 2014, Kim 2016) reported on the addition of health care support workers and relationship with outcomes. Gerova (2014) found that increases in health care assistants were not significantly related to the probability of emergency section (OR=0.99, 95%CI 0.96-1.03), instrumental birth (OR=1.003, 95%CI 0.96-1.05) or normal birth (OR=0.99, 95%CI 0.95-1.03). Kim (2016) evaluated the impact of increasing the total number of nurses, both licenced and unlicensed. As the total workforce increased, this was not significantly associated with the risk of readmission within 30 days (RR1.01, 95% CI 1.0,1.02).

Sandall (2014) found no significant differences in outcomes for increasing support worker staff in the adjusted overall analysis. Sensitivity analyses were performed in different risk groups and parity. Increasing support workers was associated with an increase in birth with bodily integrity for lower-risk women (OR 1.04) but not for higher-risk women (OR 0.96). The chances of the healthy mother outcome being met were reduced when the number of support workers increased, irrespective of parity (ORs range from 0.87 to 0.93). Support worker staffing levels were associated with a reduced healthy baby outcome (ORs range from 0.90 to 1.00 for women of different parity). When considered together, the above findings do not highlight substantial benefits or detriments for increasing support worker numbers in the workforce.

### Effects on staff delivering care

There were no published studies which reported a numeric association between staffing levels and measures of staff wellbeing in the maternity services. No studies were found relating staff retention, job satisfaction or sickness absence to staffing levels.

### Economic analyses

Economic analyses were included in primary studies by Clark (2014) and Sandall (2014). Clark (2014) noted that considerable investment would be required to implement one-to-one care for patients undergoing oxytocin induction or augmentation within the USA. They found insufficient evidence of benefit in their trial to justify the additional costs.

Sandall (2014) modelled staffing in relation to cost per birth and found that higher midwifery staffing was associated with increased delivery costs. The relationship was not strong, and this variable plus Trust size and case mix accounted for only 17% of cost variation between Trusts. Cookson et al. (2014) provided an economic impact assessment based on the Sandall (2014) data above. In their calculations, an increase in 1 Full Time Equivalent midwife per 100 births provided an incremental cost effectiveness ratio of £85,560 per additional healthy mother and £193,426 per mother with bodily integrity.

## Discussion

The body of evidence on midwifery staffing and outcomes is small and provides mixed results. While there is some evidence that increased staffing improves outcomes for mothers and neonates, this predominantly relates to labour care and outcomes within the first hour after birth. Some of the variables measured in the studies are measures of care and it is unclear whether they would translate into improved outcomes.

For the mother, increased staffing was associated with reduced epidural rates, augmentation, perineal damage during the birth, post-partum haemorrhage, and maternal readmission. For neonates, increased staffing was associated with higher breastfeeding rates and reduced need for neonatal resuscitation. Staffing may influence the quality of care in labour, as there was some evidence of improved record keeping and timeliness of emergency caesarean section. Increased attention by staff may reduce the risk of negative outcomes, while also supporting coping mechanisms in labour and supporting infant feeding.

Very few studies have suggested a negative impact of increasing staffing rates, although a large number have found no significant differences. It is possible that other prognostic variables such as patient demographics, clinical risk, or other therapeutic interventions may have overshadowed any effects of variation in staffing in these studies. A significant limitation of the available evidence is that many of the studies have not measured staffing levels directly, which has an unknown effect on the accuracy of findings. A lack of risk adjustment is a major potential source of bias within many of the studies presented.

Results for mode of birth are hard to interpret as studies are not in agreement on whether rates of spontaneous birth, instrumental birth or caesarean section are associated with staffing levels. Higher staffing levels can result from the assessed need for more staff to care for high risk mothers. This tends to mask the beneficial effect of higher staffing. Assisted birth may be entirely appropriate for high risk cases to prevent adverse maternal and fetal outcomes so should not be considered to be a detrimental outcome (Kirkup, 2015; Dietz et al., 2016).

This review contributes to the debate on whether staffing ratios should be recommended in maternity care, including all in-patient wards. It is notable that staffing ratios for labour ward, antenatal and postnatal areas have been recommended in Australia (Australian Nursing Midwifery Foundation, 2015) and in the USA (Association of Women’s Health Obstetric Neonatal Nurses, 2010). In the UK, guidance states that a systematic process is used to calculate total midwifery staff, incorporating historical data and predicted demand (National Institute for Health and Care Excellence, 2015). Birthrate Plus is one such tool for workforce planning, which is based on indicators of need in the population, while facilitating one to one care in labour (Ball et al., 2015). The tool does not collect data on outcomes, and therefore the adequacy of recommended resources cannot be evaluated. The impact of reducing or increasing staffing on outcomes is a pertinent question, especially as resources are scarce and staffing decisions should maximise cost-utility.

Understaffing may result from the inability to employ and retain registered staff (Heinen et al., 2013). This may result in the recruitment of alternative staff to complement existing midwives. This scoping review has found only three studies relating the number of support staff to patient outcomes. Outcomes were not improved by the addition of support workers, and Sandall et al. (2014) noted reductions in the composite outcome of healthy mother and healthy baby as the number of support workers increased. This fits with recent research in the nursing literature suggesting detrimental effects of diluting skill mix or having more or less nursing assistants than the average level (Aiken et al., 2017; Griffiths et al., 2019).

Makhfudli et al. (2020) found that increasing nursing staffing was associated with less risk of maternal death or severe maternal outcomes, but the same was not true for midwives. It is possible that midwives were allocated the most complex obstetric cases who had a higher background risk for poor outcomes, or that nurses had improved training in preventing escalation of potentially life threatening conditions. The skill mix of the maternity workforce is changing, and additional skills are needed to care for women and babies with complex care needs and co-morbidities. The survey by Dent (2020) found that in the UK, healthcare providers were employing a variety of personnel to support midwifery services, including theatre nurses, obstetric nurses, nursery nurses, maternity care assistants and breastfeeding supporters. The contribution of each of these staff groups towards outcomes is unclear. These initiatives may be driven by necessity rather than optimal workforce planning.

No research studies were found examining associations between staffing numbers and the wellbeing of midwives. In an online survey of almost 2000 midwives by Hunter et al. (2019), perceived inadequacy of resources was the strongest predictor of work-related burnout. This may lead to staff attrition (Heinen et al., 2013), which is costly, not only for the employer but also considering the cost of training each midwife. The State of the World’s Midwifery report highlighted voluntary attrition as one of the ten essential areas for workforce planning (Lopes et al., 2017). Challenges in recruitment and attrition have been described as a gathering storm especially in the light of increased demands and complexity (Royal College of Midwives, 2017).

It is important to note that most studies have been conducted on the labour ward/delivery suite, with a dearth of studies in antenatal and postnatal wards. Escalation plans often involve redeploying staff from these areas in order to meet need on the labour ward (Royal College of Midwives, 2016) and if they are not well staffed at the outset this may lead to critical shortages. In future, more resources may be deployed in the community as Renfrew et al. (2014) recommend a change in focus from the recognition and treatment of pathology for the minority, to providing skilled care for all. With a finite number of midwives available, this may lead to difficult choices in the distribution of staff.

### Strengths and limitations

In this scoping review, literature searching was completed in a systematic way, however, there may be undetected studies in the grey literature or in press that have not been accessed. The eligibility screening was not performed independently for all the papers, so it remains possible that some excluded papers might have been included by another reviewer. The high levels of agreement obtained on samples means that it is unlikely that this would make a substantial change to the overall number of included studies or the conclusions about the body of literature as a whole. Although major methodological issues have been discussed, the quality of the evidence has not been rigorously evaluated, which is consistent with the scoping review methodology. This means that poorer quality studies have been included, and these findings are more prone to bias.

### Recommendations for further research

Further evidence is needed so that policy makers can make informed decisions about staffing levels and configurations, and the likely impact on outcomes. High quality research is needed to clarify the direction and strength of effects. Studies should examine a range of outcomes in addition to those on labour ward. These could include maternal mental health, neonatal weight loss, jaundice, sustained breastfeeding, and neonatal readmission following discharge home. Support worker contribution and the impact on workforce wellbeing also requires further research. Improved attempts should be made to measure staffing at a ward level or individual patient level if possible. The impact of different workforce configurations and staff groups should be considered. It is important that future studies adjust for underlying risk as well as other predictive factors such as parity, gestational age, pre-existing conditions, and socioeconomic status. Economic studies could model health care costs in terms of staffing numbers, but also potential cost-savings related to intervention rates in labour, readmissions and the cost of advanced neonatal care or maternal morbidity.

## Conclusion

This scoping review has found some evidence of a positive association between staffing levels and improved outcomes for women and neonates. The evidence is not conclusive and is limited by the methodological quality of studies. Further research is needed so that service providers can predict the impact of changes to skill mix and staffing levels on a wide range of patient outcomes.

## Data Availability

Not applicable

## APPENDIX 1 : Search strategy for Medline Ovid

(adapted for other online databases using exploded MeSH headings as appropriate)

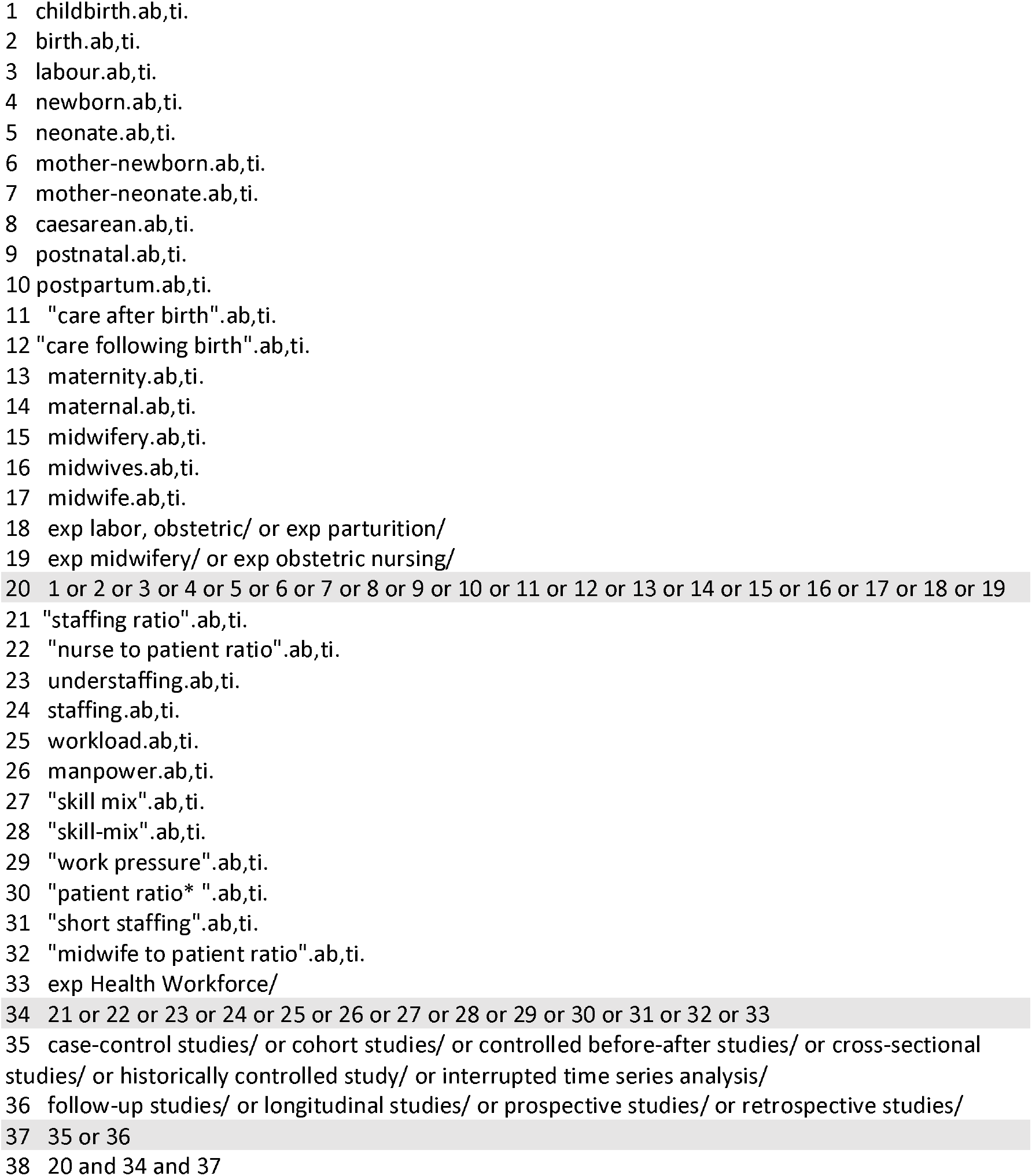

## APPENDIX 2 : Tabulation of studies

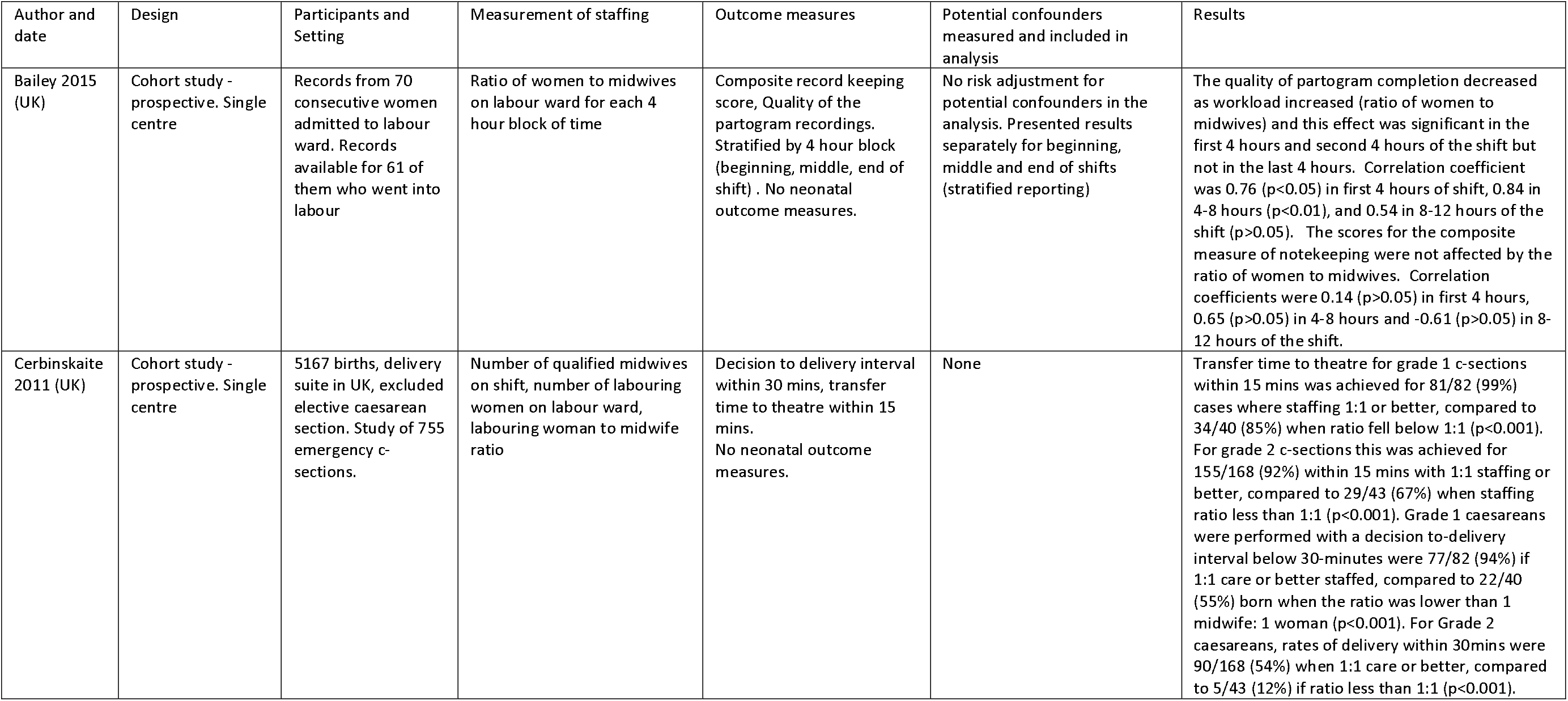
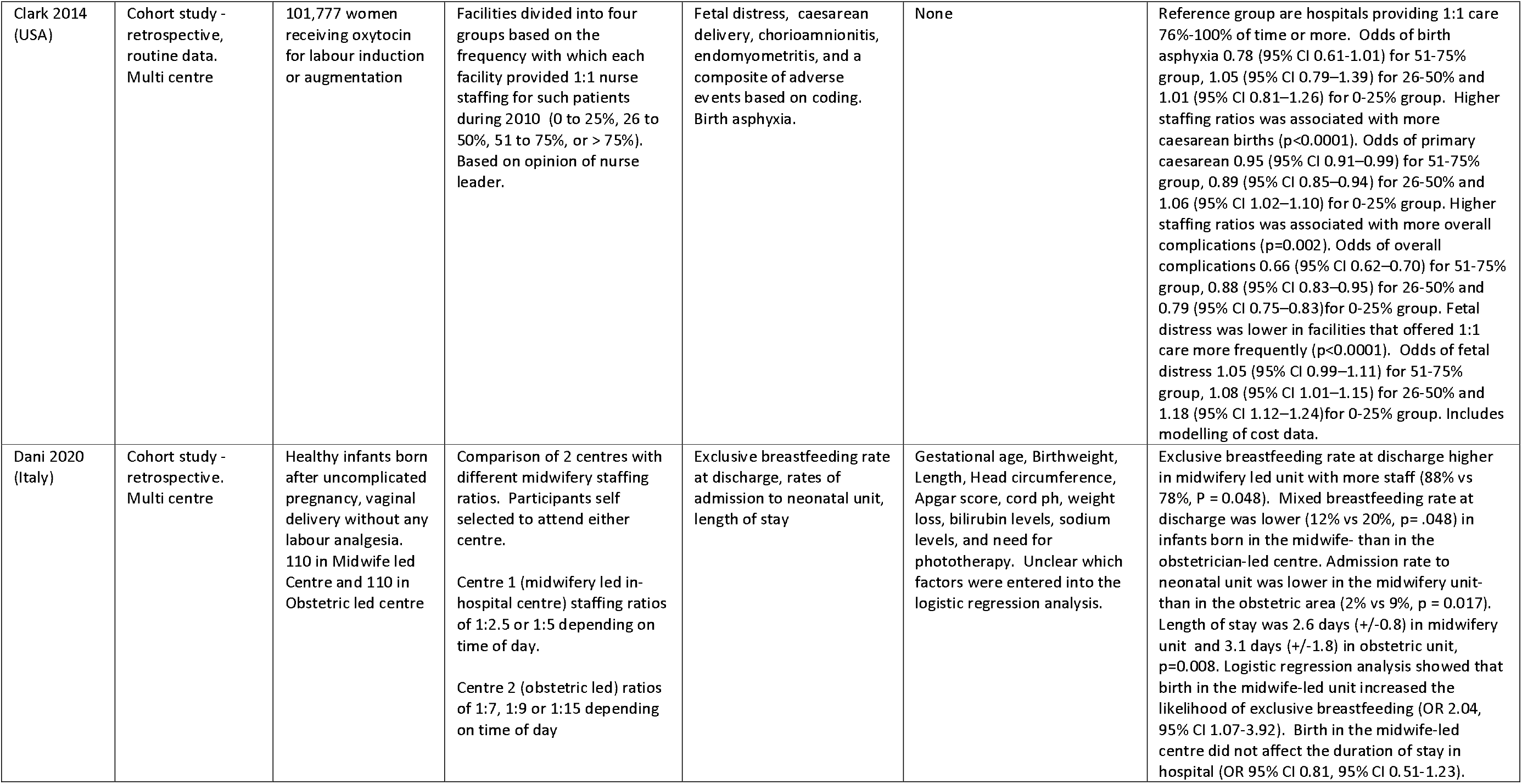
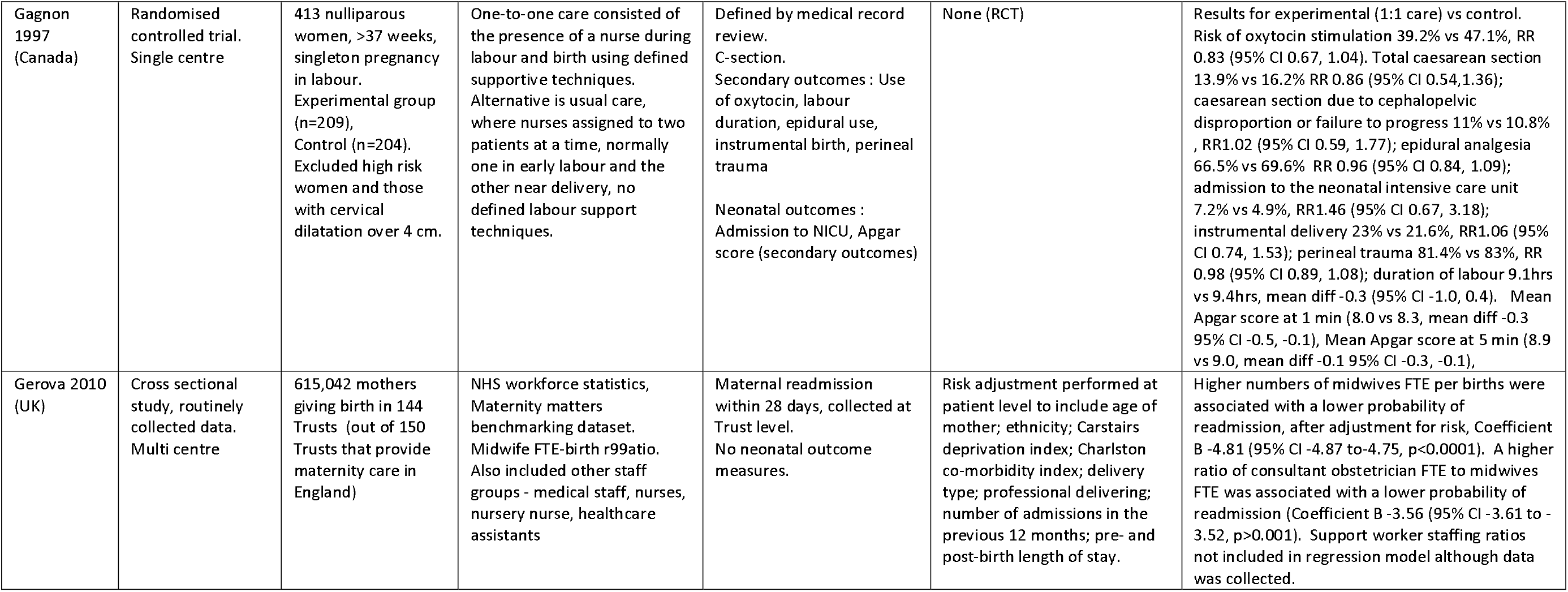
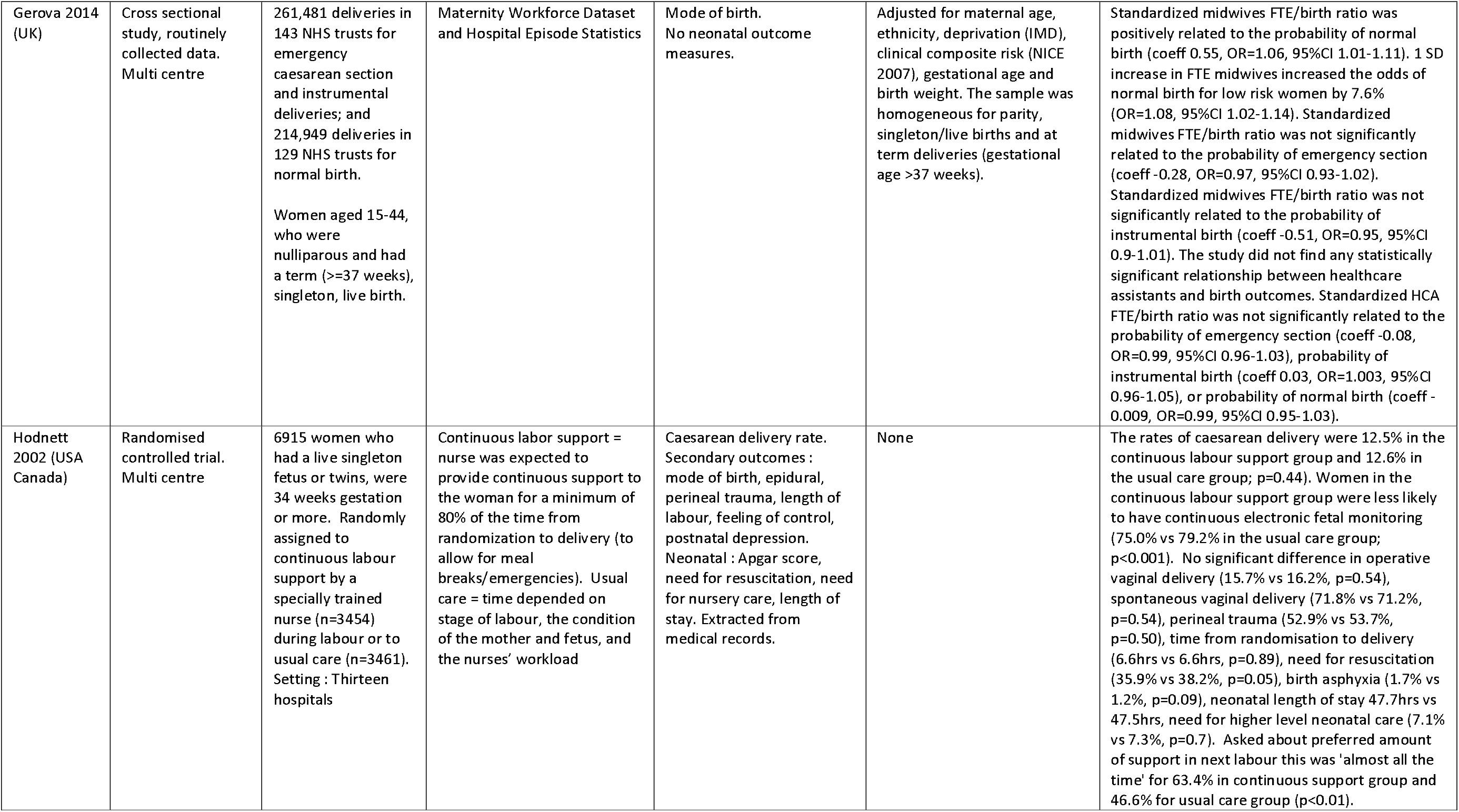
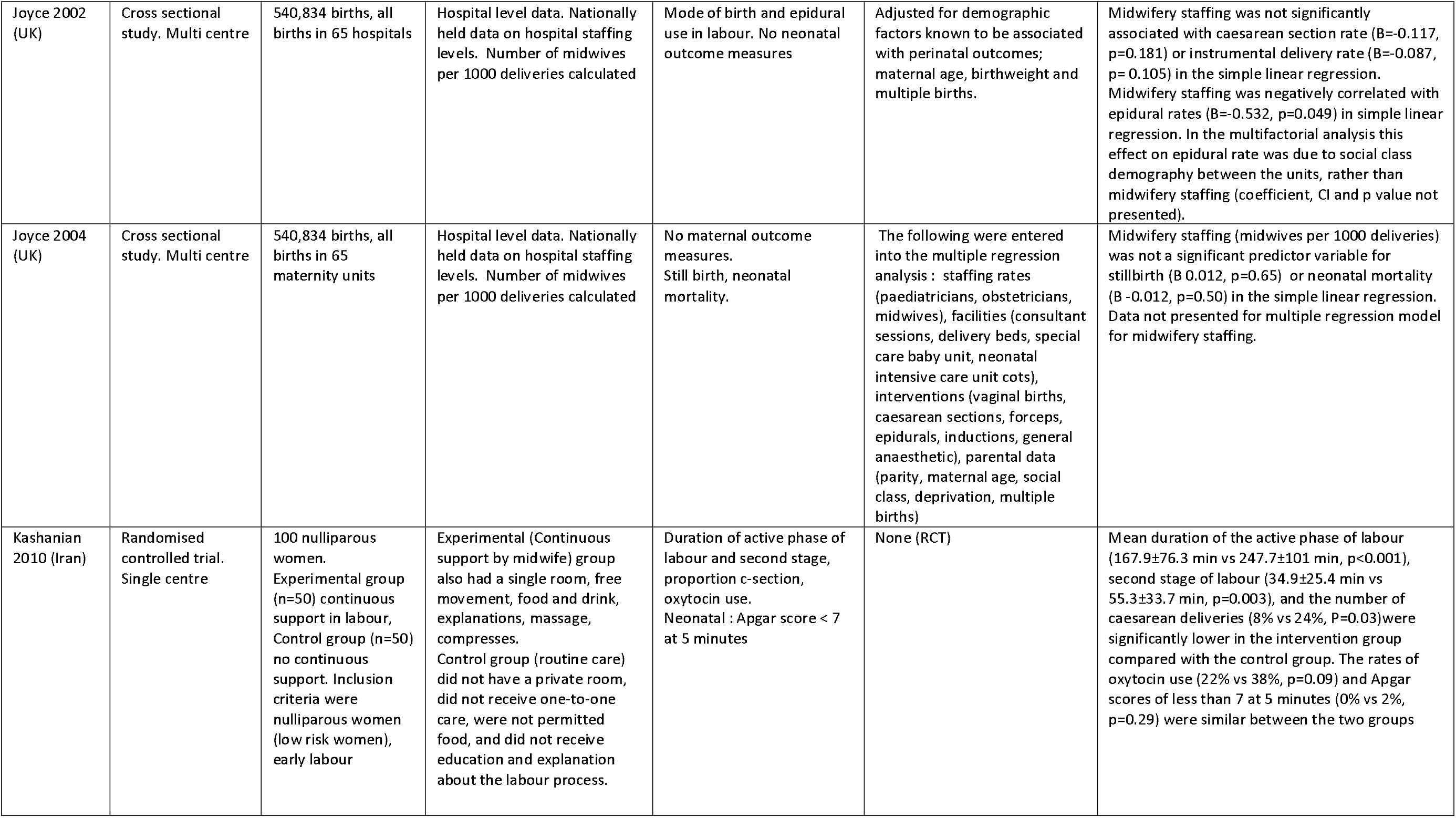
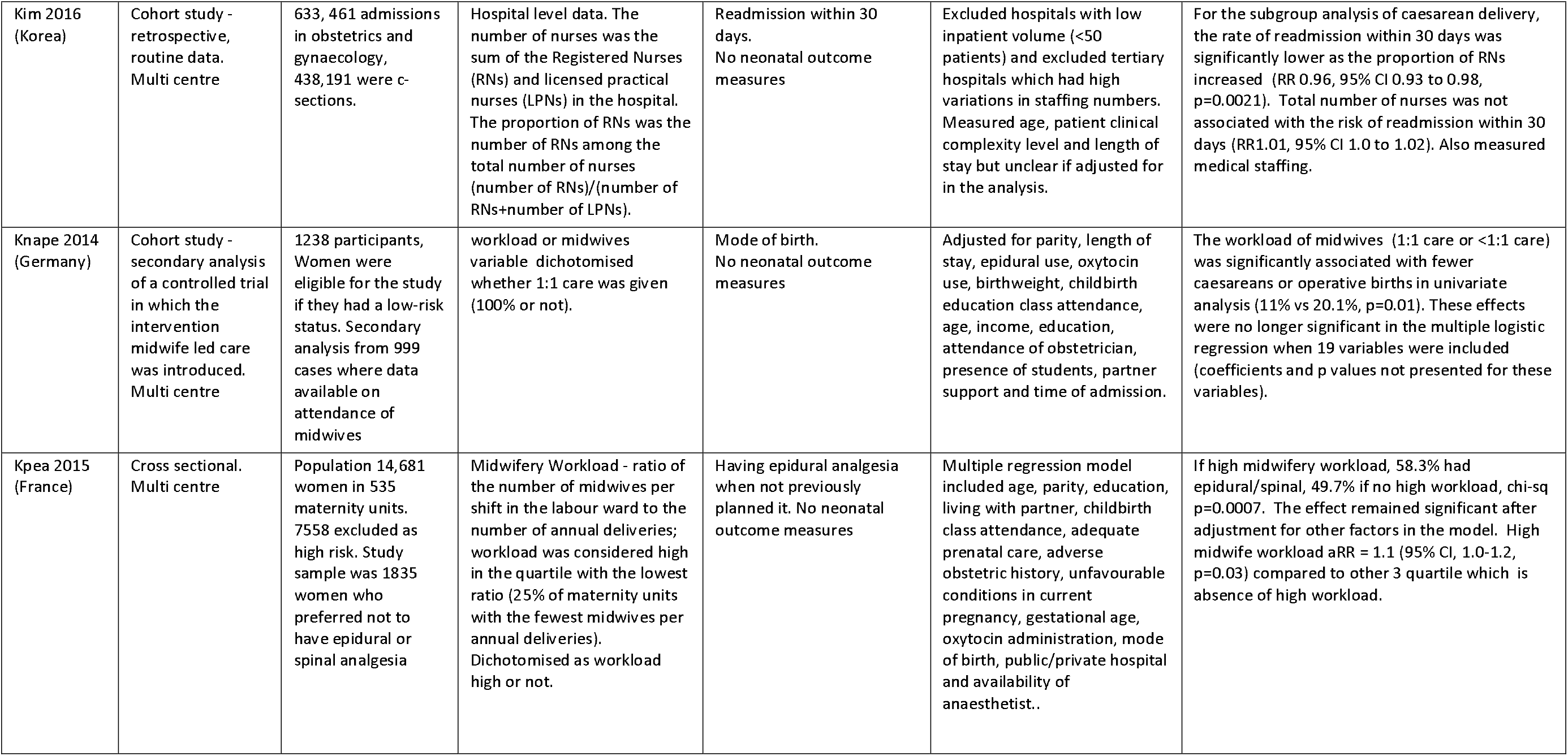
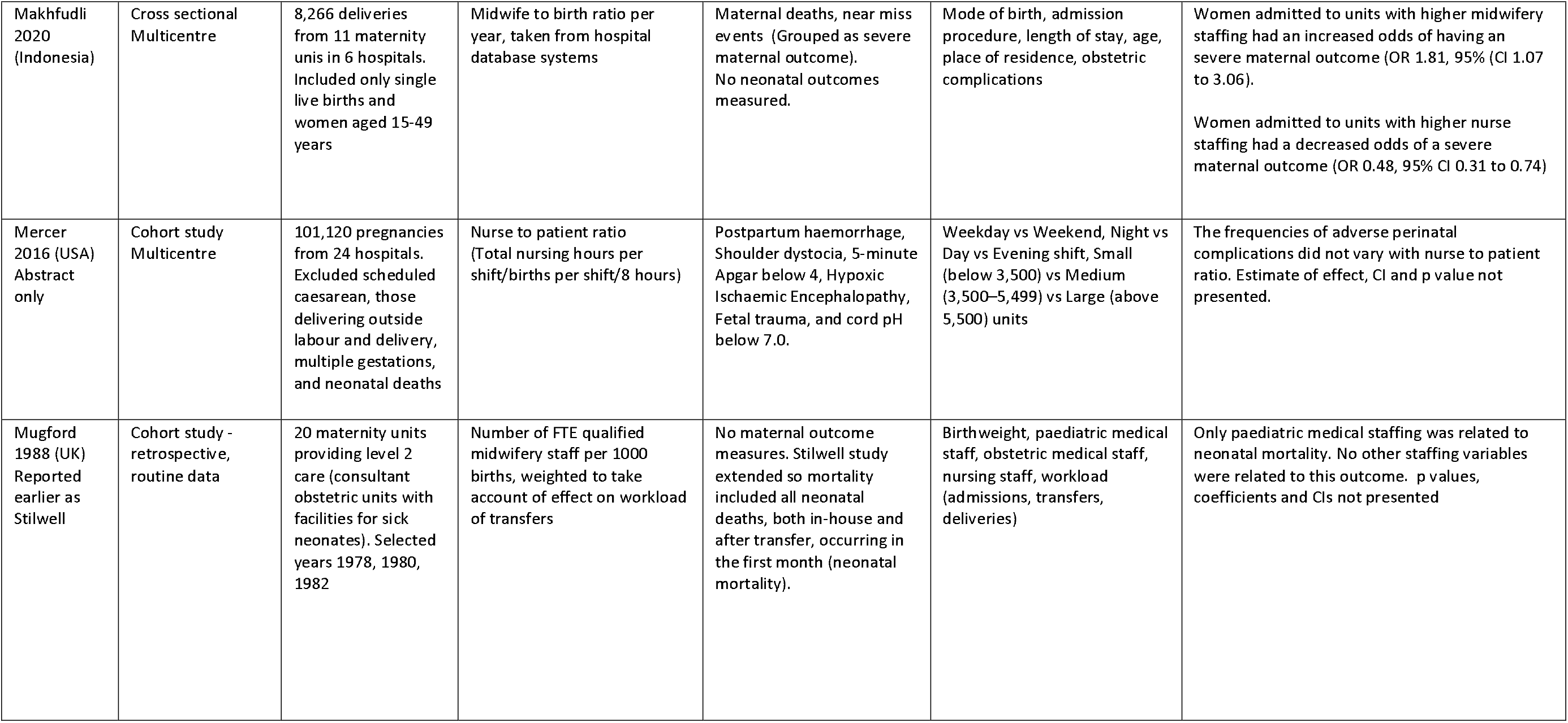
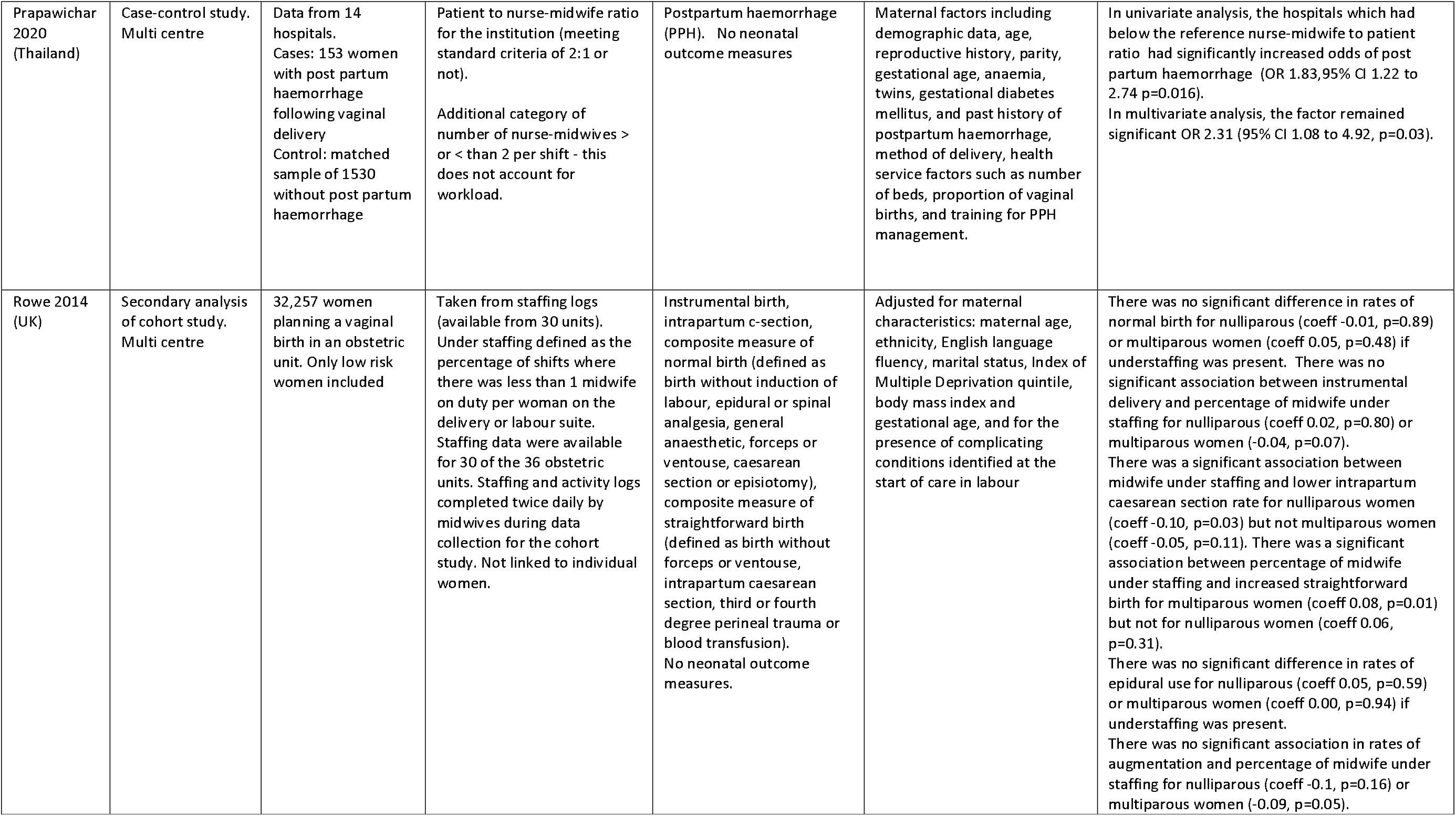
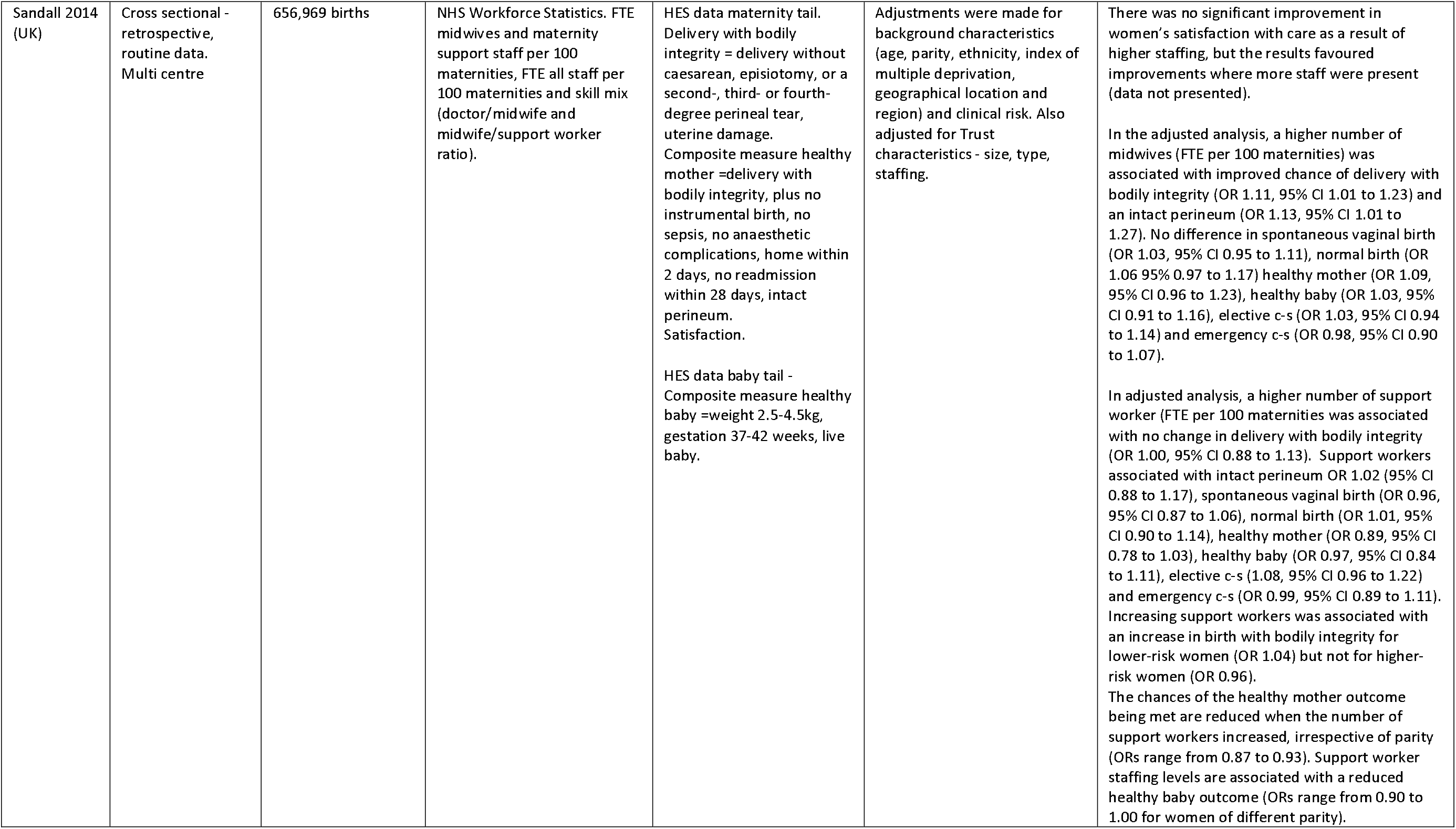
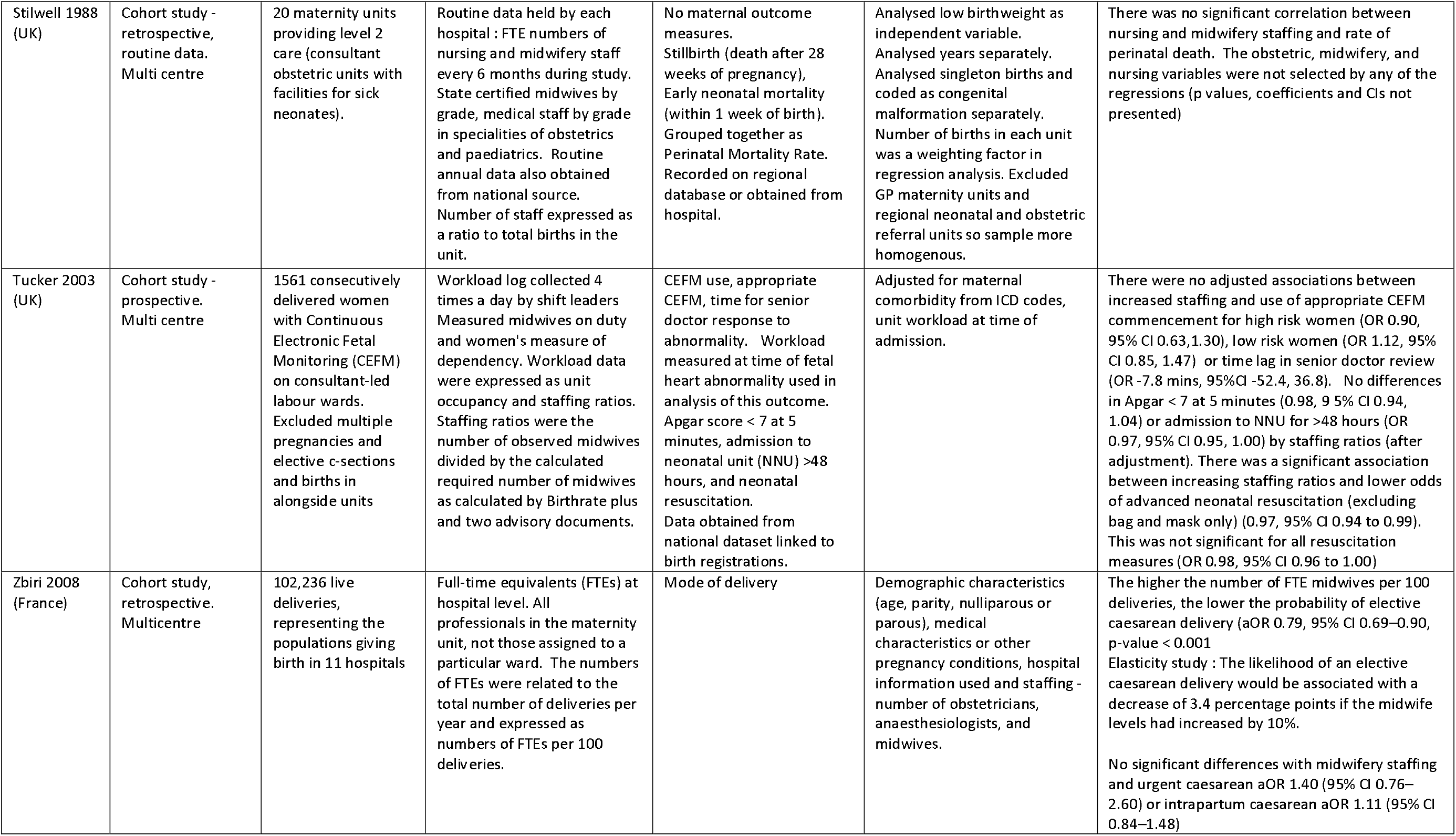

